# A digital programme to prevent falls and improve wellbeing in people living with dementia in the community: Quantitative results from the KOKU-LITE feasibility study

**DOI:** 10.64898/2026.10.27.26366422

**Authors:** Jaheeda Gangannagaripalli, Saima Ahmed, Amelia Parchment, Chunhu Shi, Schenelle Dayna Dlima, Rachel Christie, Chris Todd, Emma Vardy, Emma Stanmore

## Abstract

**Background:** Falls are common and costly among people living with Dementia (PLwD). Keep on Keep up (KOKU) is a UK National Health Service (NHS) approved tablet-based, digital gamified programme with strength and balance exercises and health literacy games, designed **with** and **for** older people at risk of falls. KOKU-LITE is a dementia-friendly accessible version of KOKU that has been co-developed **with** PLwD to meet their needs. This study aimed to assess the feasibility, usability and acceptability of KOKU-LITE among community dwelling PLwD in terms of recruitment, retention and user experience of the intervention.

**Methods:** This is a two-arm, parallel, non-blinded, feasibility randomised controlled trial. Sixty people aged ≥55 years living with dementia were recruited across Greater Manchester, UK, and randomised to either intervention group (KOKU-LITE+ dementia falls prevention leaflet) or control group (dementia falls prevention leaflet). Intervention participants were asked to use KOKU-LITE for 30 minutes a day, three times/week for six weeks. Both groups were followed up to six weeks. Primary outcomes were feasibility, usability and acceptability assessed using recruitment and retention rates, and user experience measured by user experience questionnaire. Trial registration: https://clinicaltrials.gov/study/NCT06149702.

**Findings:** Sixty participants (30 intervention, 30 control) were recruited and followed up between March 2024 to September 2025. Among those approached, 87% (60/69) consented to participate, 90% (54/60) were retained at six weeks follow up. Overall, the user experience findings indicated strong acceptability and generally positive perceived usability and engagement. No serious adverse events, including injurious falls, were reported during the intervention period.

**Interpretation:** KOKU-LITE was feasible to deliver and acceptable to PLwD in a home-based community setting. Although the study was not powered to assess effectiveness, the findings support progression to a definitive trial, with refinement of outcome selection, intervention-use monitoring and follow-up duration.

**Funding:** National Institute for Health and Care Research (NIHR) and Alzheimer’s Society (Award number: NIHR200174).

## 1. INTRODUCTION

Dementia is a major public health challenge in the UK and globally. It is estimated that approximately 982,000 people are currently living with dementia in the UK, and this figure is projected to increase to 1.4 million by 2040^(1)^. The costs of dementia in the UK, including healthcare, social care, unpaid care, reduced quality of life and economic losses, is estimated to be around £42 billion in 2024, and this figure is projected to exceed £90 billion by 2040^(1)^. Dementia has been recognised by the World Health Organization (WHO) as a global public health priority due to its increasing prevalence and its substantial impact on people living with dementia (PLwD), their families, carers, health systems, and wider society ^(2)^. Dementia is the seventh leading cause of death worldwide and one of the leading causes of disability and dependency among older people globally^(3)^. Dementia is also one of the leading causes of death in the UK^(4)^.

Falls are a major global and UK public health concern and the second leading cause of unintentional injury-related mortality worldwide ^(5)^. Globally, falls account for approximately 684,000 deaths and 172 million fall-related disabilities annually ^(6)^. In the UK, around one-third of people aged ≥65 years fall each year, while fragility fractures are estimated to cost £4.4 billion annually, with hip fractures alone accounting for around £2 billion ^(7,8)^. PLwD are more than twice as likely to fall; with a meta-analysis estimating an annual prevalence of 44.27% and an average of 1.30 falls per person-year ^(9,10)^. However, dementia-specific falls prevention evidence remains limited, insufficient and inconclusive ^(11–13)^ to support routine implementation across community and care-home settings.

Digital technologies are increasingly promoted to support health and wellbeing, including by the WHO^(14)^. For PLwD, digital health interventions may support independence, reduce falls, improve quality of life, and lower health and social care costs. However, robust evidence on effective digital technologies for PLwD remains limited. *Eost-Telling et.al*, in their systematic review, found lack of evidence on the use of apps for falls prevention in PLwD^(15)^. While feasibility studies suggest potential, they are limited by small samples and the absence of control groups^(16)^. Adoption is further constrained by dementia progression, limited digital literacy, anxiety, cognitive fatigue, and usability barriers^(17)^. Digital interventions therefore require careful adaptation to ensure they are acceptable and usable for PLwD.

Keep-On-Keep-Up (KOKU)^(18,19)^ is an NHS-approved digital gamified programme that includes strength and balance exercises with health literacy games (including home safety, hydration, nutrition, bone and brain health) and has been co-designed **with** and **for** older adults at risk of falls. KOKU-LITE is a dementia friendly accessible version of KOKU that has been co-developed **with** PLwD and carers of PLwD. Given the limited evidence for digital falls-prevention interventions in this population, this study evaluates the feasibility, acceptability, and usability of KOKU-LITE among PLwD in terms of recruitment, retention and user experience of the intervention.

## 2. METHODS

This feasibility randomised controlled trial (fRCT) is reported in accordance with CONSORT 2010 (online supplementary appendix 1) and the Template for Intervention Description and Replication (TIDieR) guidance (online supplementary appendix 2). Full methodological details are available in the published study protocol ^(20)^.

### Study design and setting

We conducted a two-arm, parallel-group, non-blinded, six-week fRCT with an embedded mixed-methods evaluation. The trial compared the intervention (KOKU-LITE+ dementia specific falls prevention leaflet) with control (dementia specific falls prevention leaflet) among community-dwelling PLwD in Greater Manchester, UK.

A project advisory group (n=4) comprising of PLwD (n=2; Male=1, a person with vascular dementia; Female=1, a person with Alzheimer’s disease) and carers of PLwD (n=2;Male=1, carer for a person with Alzheimer’s disease; Female=1, carer for a person with vascular dementia) provided feedback to the team throughout the study including design, recruitment, interpretation, and dissemination of the study.

### Participants and recruitment

PLwD were eligible if they were aged ≥55 years, had any type but varying levels of dementia from mild to moderate stages (ratings 1 and 2 on clinical dementia rating (CDR) scale^(21)^), able to provide consent, able to speak English, and mobile (See Table 1 for the full eligibility criteria).

**Table 1.**
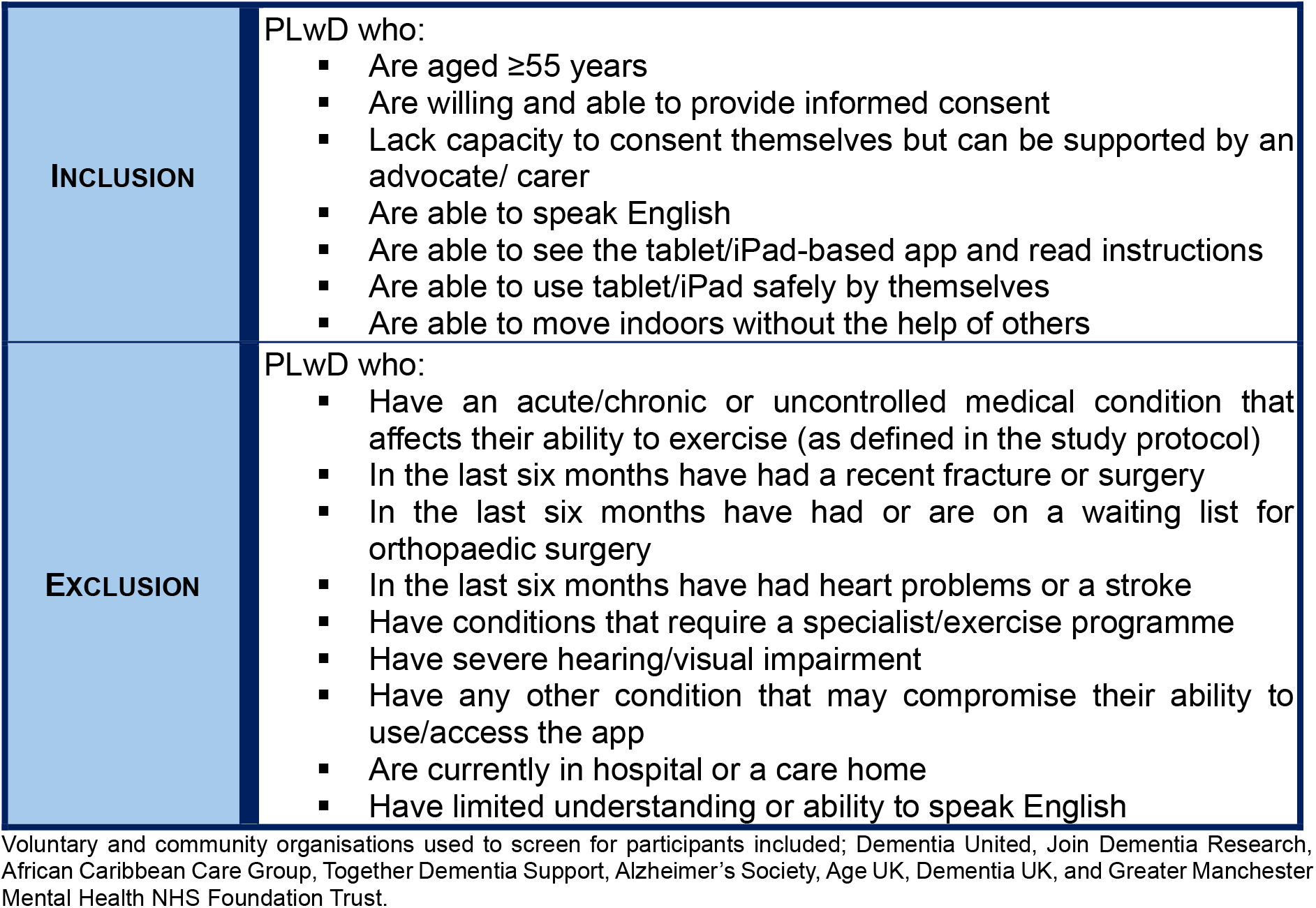
Study participant inclusion and exclusion criteria.

|  |  |
| --- | --- |
| **INCLUSION** | PLwD who:<br>▪ Are aged $\geq 55$ years<br>▪ Are willing and able to provide informed consent<br>▪ Lack capacity to consent themselves but can be supported by an advocate/ carer<br>▪ Are able to speak English<br>▪ Are able to see the tablet/iPad-based app and read instructions<br>▪ Are able to use tablet/iPad safely by themselves<br>▪ Are able to move indoors without the help of others |
| **EXCLUSION** | PLwD who:<br>▪ Have an acute/chronic or uncontrolled medical condition that affects their ability to exercise (as defined in the study protocol)<br>▪ In the last six months have had a recent fracture or surgery<br>▪ In the last six months have had or are on a waiting list for orthopaedic surgery<br>▪ In the last six months have had heart problems or a stroke<br>▪ Have conditions that require a specialist/exercise programme<br>▪ Have severe hearing/visual impairment<br>▪ Have any other condition that may compromise their ability to use/access the app<br>▪ Are currently in hospital or a care home<br>▪ Have limited understanding or ability to speak English |

Voluntary and community organisations used to screen for participants included; Dementia United, Join Dementia Research, African Caribbean Care Group, Together Dementia Support, Alzheimer's Society, Age UK, Dementia UK, and Greater Manchester Mental Health NHS Foundation Trust.

We used several strategies to recruit participants into the study: 1) Participants were recruited through voluntary and community organisations across Greater Manchester (Table 1). 2) Snowball sampling was also used, with participants encouraged to share study information with other potentially eligible PLwD. 3) And the research team also presented the study at local community events, where interested individuals completed consent-to-contact forms. Potential participants were contacted by members of the research team (JG, SA, and AP) to assess their eligibility and enrol them into the study. The intended sample size for this feasibility study is 60 PLwD allocated 30 per group, allocated 1:1 by simple randomisation (further details provided in the protocol paper ^(20)^).

### Qualitative sample

A purposive sample of 15 PLwD and/or carersof PLwD who had consented to participate in the fRCT were invited to take part in post-intervention semi-structured interviews exploring the feasibility of the study, usability and acceptability of KOKU-LITE. Health and social care professionals were recruited through personal and professional networks in the UK, including, Join Dementia Research, Alzheimer’s Society, Age UK, Manchester University NHS Foundation Trust, Tameside & Glossop Integrated Care NHS Foundation Trust, Northern Care Alliance NHS Foundation Trust and Greater Manchester Mental Health NHS Foundation Trust. Two focus groups were conducted with 11 professionals, including Doctors, Consultant Geriatricians, Hospital and Community Nurses, Physiotherapists, Occupational Therapists, support workers, dementia advisors and PPIE representatives to explore the usability, acceptability, and motivations for using KOKU-LITE with PLwD in the community. Qualitative findings will be reported separately in a future paper.

### Consent

All potential participants received written study information and were given at least one week to decide whether to take part in the study. Written or audio-recorded informed consent was obtained from all participants and/or their carers.

### Randomisation and masking

After baseline assessment, eligible participants were randomly assigned to the intervention or control group by an independent researcher (CS), who was not involved in data collection. Allocation used permuted block randomisation with randomly varying block sizes of four and six, implemented through Sealed Envelope ^(22)^. Because of the nature of the intervention, participants, researchers, and outcome assessors could not be masked to treatment allocation. Participants were informed of their group allocation after baseline data had been collected.

### Intervention and comparator

KOKU-LITE (accessed from KOKU Health website: https://kokuhealth.com/) is an NHS-approved digital gamified strength and balance exercise programme with health literacy games (including home hazards, hydration, nutrition, bone and brain health) informed by the principles of the evidence-based FAME and Otago exercise programmes^(23,24)^. During set-up, users completed a pre-screening questionnaire, and the programme was tailored to their needs and functional ability. The graded exercise schedule began with low intensity exercises and progressed to more challenging exercises.

Intervention was delivered by the trained and experienced researchers (JG, AP, SA) in participants’ homes across Greater Manchester, UK. Participants allocated to the intervention group were provided with an iPad with KOKU-LITE installed, a short instruction booklet and dementia-specific falls prevention leaflet which they followed over a period of six weeks. They were advised to use KOKU-LITE for 30 minutes per session, three times per week. Participants attended a 60-minute training session, delivered either in small groups of two to four participants or one-to-one, depending on preference and support needs; carers attended where required. During the intervention period, researchers contacted participants by telephone to support adherence and troubleshoot technical issues. Intervention participants were also encouraged to read and use the dementia specific falls prevention leaflet. Participants allocated to the control group were advised to read and follow the dementia specific falls prevention leaflet and were offered time-limited access to KOKU-LITE after completion of the six-week follow-up.

### Outcomes and data collection

Outcome measures were collected at baseline and six weeks by the research team for both trial groups. Demographic and clinical data were collected at baseline and included sex, age, ethnicity, country of birth, marital status, socioeconomic status, dementia diagnosis and severity, computer use, vision, medical history, history of falls and fractures, medication use, and long-term conditions affecting ability to exercise.

### Primary outcomes

The feasibility, of KOKU-LITE was reported using recruitment and retention rates, while usability, and acceptability measured using the User Experience Questionnaire (UEQ). The feasibility and acceptability of the intervention will be evaluated in terms of recruitment, retention and adherence rates throughout the trial in accordance with the progression criteria (accessed from the protocol paper^(20)^).

### Secondary outcomes

The outcomes including quality of life, participants’ activities of daily living (ADL) functional ability, lower limb strength, concerns of falling, falls risk and mood were assessed using standardised instruments and assessments, including European Quality of Life Healthy Questionnaire (EQ-5D-5L)^(25)^, Erlangen Test for Activities of Daily Living (E-ADL)^(26)^, Short Physical Performance Battery (SPPB)^(27)^, Fall Risk Assessment Tool (FRAT)^(28)^, Short Falls Efficacy Scale International (Short FES-I)^(29)^, and 5-item Geriatric Depression Scale (5-GDS)^(30)^ respectively. Outcome measures and cut-off scores used are summarised in Table 2.

**Table 2:** Study outcome measures and cut-off scores.

| OUTCOME MEASURE | SCORE FORMAT | SCORE INTERPRETATION |
| --- | --- | --- |
| **Erlangen Activities of Daily Living (E-ADL)** | Score I<br>Score II<br>Score III<br>Score IV<br>Score V | TOTAL of all 5 sub-scores.<br>○ 0-9 Points = reflects profound functional impairment and high dependency<br>○ 10-17 = indicates clear decline in functional independence, requires assistance in many areas<br>○ 18-23 = early stages of dementia, performance in complex daily activities begins to decline<br>○ 24-30 = indicates high function/ minimal impairment, with potential MCI, but no clear dementia |
| **Short Physical Performance Battery (SPPB)** | Balance test score<br>Gait speed test score<br>Repeated chair stand test score | TOTAL of all 3 sub-scores.<br>○ ≤8 points = recommended screening for sarcopenia<br>○ ≤9 points = suggests frailty and disability risk<br>○ ≤10 points = increased risk of future disability<br>○ =10-12 points = represents high physical performance |
| **Fall Risk Assessment Tool (FRAT)** | Yes or no responses to 5 questions | TOTAL number of yes responses.<br>○ <3 yes = low risk<br>○ 3-5 yes = high risk |
| **Short Falls Efficacy Scale (Short FES-1)** | Not at all concerned (1)<br>Somewhat concerned (2)<br>Fairly concerned (3)<br>Very concerned (4) | TOTAL score of all responses.<br>○ 7-8 = low concern<br>○ 9-13 = moderate concern<br>○ 14-28 = high concern |
| **European Quality of Life Healthy Questionnaire (EQ-5D-5L)** | No problems (1)<br>Slight problems (2)<br>Moderate problems (3)<br>Severe problems (4)<br>Unable/ extreme problems (5) | TOTAL score of all 5-dimension responses.<br>○ 11111: no health problems (perfect health)<br>○ 55555: extreme problems in all dimensions (worst health) |
| **5-item Geriatric Depression Scale (5-GDS)** | Q1 response no (1)<br>Q2 response yes (1)<br>Q3 response yes (1)<br>Q4 response yes (1)<br>Q5 response yes (1) | TOTAL scores of highlighted responses.<br>○ 0-1 = not depressed<br>○ 2+ = depressed |

Assessments were conducted face-to-face either in participants’ homes or at dementia cafés, depending on participant preference and convenience. Where possible, the same member of the research team completed baseline and follow-up assessments for each participant as some participants reported feeling more comfortable with this continuity. Follow up outcome data were collected even when participants discontinued use of KOKU-LITE during the intervention period.

### User experience with KOKU-LITE

Participants allocated to KOKU-LITE completed the UEQ^(31)^ at six-week follow-up. UEQ findings were interpreted alongside qualitative interview data to inform understanding of perceived usability and acceptability of KOKU-LITE; these findings will be reported separately.

### Statistical analysis

Data were processed in Microsoft Excel and analysed using SPSS version 28.

The analyses were descriptive and exploratory, consistent with the feasibility study design. Baseline and six-week outcome measures were described by group and time point. For continuous outcomes, mean change scores, mean differences, standard deviations, and 95% confidence intervals were calculated to estimate the direction and precision of change and help inform the design of a future definitive trial. Categorical outcomes were summarised using frequencies and percentages. The findings of UEQ were reported descriptively to assess user experience of the digital intervention. No formal hypothesis testing was undertaken, as the study was not designed or powered to assess intervention effectiveness.

### Ethical approval

Ethical approval was granted by the Yorkshire and The Humber—Bradford Leeds Research Ethics Committee, UK (REC reference: 23/YH/0262).

## RESULTS

### Baseline characteristics

Baseline participant characteristics for all the 60 participants enrolled in the study are shown in Table 3. The mean age was 78.3 years (SD 8.0), and more than half of the participants (35 [58.3%] were male. Most participants were White British or Irish (46 [76.7%]) and were born in the UK (46 [76.7%]). Dementia type varied across the sample; Alzheimer’s disease was the most frequently reported diagnosis (21 [35.0%]), followed by mixed dementia (16 [26.7%]) and vascular dementia (9 [15.0%]). Dementia severity was most commonly reported as mild (24 [40.0%]) or moderate (17 [28.3%]), with 18 participants (30.0%) unsure of dementia stage. Baseline characteristics were summarised descriptively; no statistical tests were used to compare groups at baseline.

**Table 3:** Participant characteristics at baseline.

| **Participant details**<br>n (%) or mean±SD | **Total**<br>(n=60) | **KOKU-LITE**<br>(n=25) | **Control**<br>(n=29) | **Withdrawals**<br>(n=6) |
| --- | --- | --- | --- | --- |
| **Age - years** (mean±SD) | 78.3±8.0 | 76±9.7 | 80±6.3 | 3 missing entries |
| **Sex** |  |  |  |  |
| Male | 35 (58.3) | 17 (68.0) | 13 (44.8) | 5 dropouts |
| Female | 25 (41.7) | 8 (32.0) | 16 (55.2) | 1 dropout |
| **Ethnicity** |  |  |  | *1 missing entry |
| Caribbean | 2 (3.3) | 0 | 2 (7.1) | - |
| African | 3 (5.0) | 2 (8.0) | 1 (3.6) | - |
| White British | 45 (75.0) | 18 (72.0) | 22 (78.6) | 5 dropouts |
| Irish | 1 (1.7) | 1 (4.0) | 0 | - |
| Other white | 2 (3.3) | 2 (8.0) | 0 | - |
| Indian | 1 (1.7) | 1 (4.0) | 0 | - |
| Pakistani | 4 (6.7) | 1 (4.0) | 3 (10.7) | - |
| Other ethnic | 1 (1.7) | 0 | 0 | 1 dropout |
| **Birth country** |  |  |  | *1 missing entry |
| UK | 46 (76.7) | 19 (76.0) | 22 (78.6) | 5 dropouts |
| Ireland | 2 (3.3) | 1 (4.0) | 1 (3.6) | - |
| Caribbean | 1 (1.7) | 0 | 1 (3.6) | - |
| East Africa | 3 (5.0) | 2 (8.0) | 1 (3.6) | - |
| West Africa | 1 (1.7) | 0 | 1 (3.6) | - |
| Middle East | 1 (1.7) | 0 | 0 | 1 dropout |
| India | 2 (3.3) | 1 (4.0) | 1 (3.6) | - |
| Other | 3 (5.0) | 2 (8.0) | 1 (3.6) | - |
| **Marital status** |  |  |  | *1 missing entry |
| Single | 3 (5.0) | 2 (8.0) | 1 (3.6) | - |
| Married/ Partner | 41 (68.3) | 17 (68.0) | 19 (67.9) | 5 dropouts |
| Divorced | 3 (5.0) | 2 (8.0) | 0 | 1 dropout |
| Separated | 1 (1.7) | 1 (4.0) | 0 | - |
| Widowed | 11 (18.3) | 3 (12.0) | 8 (28.6) | - |
| **Work status** |  |  |  | 2 missing entries |
| Retired | 57 (95.0) | 23 (95.8) | 28 (100.0) | 6 dropouts |
| Unemployed | 1 (1.7) | 1 (4.2) | 0 | - |
| **Paid job (yes)** | 59 (98.3) | 25 (100.) | 28 (100.0) | 6 dropouts |
| **Work position** |  |  |  | *1 missing entry |
| Self-employed (25+) | 2 (3.3) | 2 (8.0) | 0 | - |
| Self-employed (<25) | 6 (10.0) | 4 (16.0) | 2 (7.1) | - |
| Self-employed (0) | 4 (6.7) | 1 (4.0) | 2 (7.1) | 1 dropouts |
| Manager (25+) | 9 (15.0) | 4 (16.0) | 4 (14.3) | 1 dropouts |
| Manager (<25) | 5 (8.3) | 0 | 4 (14.3) | 1 dropouts |
| Supervisor | 9 (15.0) | 6 (24.0) | 2 (7.1) | 1 dropouts |
| Employee | 24 (40.0) | 8 (32.0) | 14 (50.0) | 2 dropouts |
| **Technology use (yes)** |  |  |  | - |
| Own tablet computer | 27 (45.0) | 8 (32.0) | 16 (55.2) | 3 dropouts |
| Need computer help | 23 (38.3) | 6 (24.0) | 14 (48.3) | 3 dropouts |
| Own smartphone | 41 (68.3) | 14 (56.0) | 24 (82.8) | 3 dropouts |
| Need smartphone help | 23 (38.3) | 8 (32.0) | 13 (44.8) | 2 dropouts |
| Own laptop | 25 (41.7) | 10 (40.0) | 13 (44.8) | 2 dropouts |
| Need laptop help | 25 (41.7) | 9 (36.0) | 14 (48.3) | 2 dropouts |
| **Person type** |  |  |  |  |
| PLwD | 35 (58.3) | 19 (76.0) | 13 (44.8) | 3 dropouts |
| Relative/ friend | 24 (40.0) | 5 (20.0) | 16 (55.2) | 3 dropouts |
| Carer of PLwD | 1 (1.7) | 1 (4.0) | 0 | - |
| **Dementia type** |  |  |  | *1 missing entry |
| Alzheimer's | 21 (35.0) | 13 (52.0) | 6 (20.7) | 2 dropouts |
| Lewy Body | 2 (3.3) | 0 | 2 (6.9) | - |
| Vascular | 9 (15.0) | 2 (8.0) | 7 (24.1) | - |
| Frontotemporal | 1 (1.7) | 0 | 1 (3.4) | - |
| Mixed | 16 (26.7) | 7 (28.0) | 7 (24.1) | 2 dropouts |
| Do not know | 10 (16.7) | 3 (12.0) | 6 (20.7) | 1 dropout |
| **Dementia stage** |  |  |  | *1 missing entry |
| Mild | 24 (40.0) | 8 (33.3) | 14 (48.3) | 2 dropouts |
| Moderate | 17 (28.3) | 9 (37.5) | 7 (24.1) | 1 dropout |
| Do not know | 18 (30.0) | 7 (29.2) | 8 (27.6) | 3 dropouts |
| **Medications (yes)** | 59 (98.3) | 24 (100.0) | 29 (100.0) | 1 missing entry<br>6 dropouts |
| **Medications – number taken (mean±SD)** | 7.15±4.805 | 6.64±4.282 | 7.44±5.47 | 7 missing entries<br>6 dropouts |
| **Stroke 6M (yes)** | 1 (1.7) | 1 (4.0) | 0 | - |
| **Heart attack 6M (yes)** | 2 (3.3) | 2 (8.0) | 0 | - |
| **Bone fracture (yes)** | 26 (43.3) | 12 (48.0) | 12 (41.4) | 2 dropouts |
| **Surgery (yes)** | 10 (16.7) | 3 (12.0) | 4 (13.8) | 3 dropouts |
| **Joint replacement (yes)** | 14 (23.3) | 7 (28.0) | 6 (20.7) | 1 dropout |
| **Eyesight** |  |  |  |  |
| Excellent | 5 (8.3) | 2 (8.0) | 3 (10.3) | - |
| Good | 31 (51.7) | 13 (52.0) | 13 (44.8) | 5 dropouts |
| Fair | 19 (31.7) | 7 (28.0) | 11 (37.9) | 1 dropout |
| Poor | 5 (8.3) | 3 (12.0) | 2 (6.9) | 0 |
| **Falls 12M** |  |  |  |  |
| Never | 24 (40.0) | 7 (28.0) | 16 (55.2) | 1 dropout |
| Once | 17 (28.3) | 10 (40.0) | 4 (13.8) | 3 dropouts |
| Twice or more | 19 (31.7) | 8 (32.0) | 9 (31.0) | 2 dropouts |

Wk=Week; M=Month; Sig.= Significance; MD= Mean Difference; SMD= Standardised Mean Difference; OR= Odds Ratio; CI= Confidence Interval; SD= Standard Deviation. \* Demographic data were partially missing for one participant.

### Outcomes

#### Primary outcomes: Recruitment, retention and user experience of the intervention

Between March 2024 and September 2025, 69 people living with dementia were screened, of whom 60 were recruited and randomly assigned to either intervention (n=30) or control (n=30), giving a recruitment rate of 87%. At six week follow up, 54 participants completed assessments: 25 in the intervention group and 29 in the control group, resulting in a retention rate of 90%. Six participants withdrew or were lost to follow-up because of the following reasons: reported family problems (n=2), death of the PLwD (n=1), hospitalisation due to stroke (n=1), severe hearing impairment (n=1), and inability to follow instructions due to progression of dementia (n=1). Demographic data were partially missing for one participant. No serious adverse events, including injurious falls, were reported during the intervention period. Recruitment and retention are summarised in Figure 1

**Figure 1:**
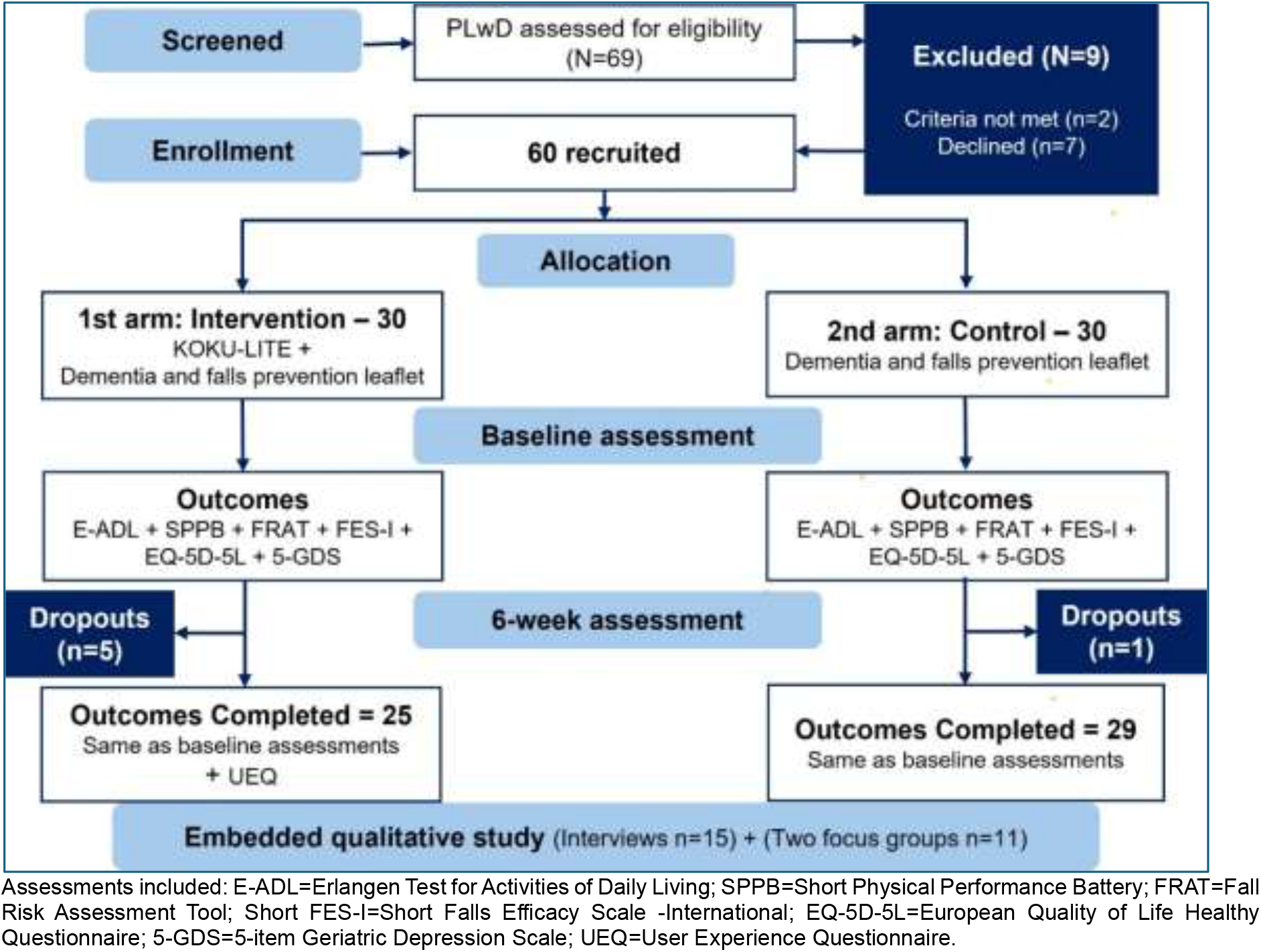
CONSORT flow diagram.

### User experience

Participants in the intervention group (24/25) also completed the UEQ at the six-week follow-up (Table 4). Perceived usability was favourable: perspicuity was rated positively (Mean = 1.78, SD = 1.24, 95% CI [1.28, 2.27]), as were efficiency (Mean = 1.17, SD = 0.97, 95% CI [0.78, 1.55]) and dependability (Mean = 1.31, SD = 0.96, 95% CI [0.93, 1.70]). Acceptability, was particularly strong (Mean = 1.92, SD = 0.64, 95% CI [1.66, 2.17]) and was classified as excellent relative to the UEQ benchmark, placing it among the top 10% of benchmarked results. Engagement was also favourable for stimulation (Mean = 1.34, SD = 1.02, 95% CI [0.94, 1.75]); however, novelty was lower (Mean = 0.77, SD = 1.09, 95% CI [0.33, 1.21]) and fell within the UEQ neutral range, despite an above average benchmark classification. Overall, the findings indicate strong acceptability and generally positive perceived usability and engagement, while perceived innovativeness represents the clearest opportunity for improvement.

**Table 4:** User Experience Questionnaire (UEQ) scores for the intervention group.

| Scale | Mean | Std. Dev. | N | Confidence | Confidence interval |  |
| --- | --- | --- | --- | --- | --- | --- |
| Attractiveness | 1.917 | 0.645 | 24 | 0.258 | 1.659 | 2.175 |
| Perspiciuity | 1.778 | 1.237 | 24 | 0.495 | 1.283 | 2.273 |
| Efficiency | 1.167 | 0.969 | 24 | 0.388 | 0.779 | 1.554 |
| Dependability | 1.313 | 0.956 | 24 | 0.383 | 0.930 | 1.695 |
| Stimulation | 1.344 | 1.016 | 24 | 0.406 | 0.937 | 1.750 |
| Novelty | 0.771 | 1.093 | 24 | 0.437 | 0.333 | 1.208 |

### Secondary outcomes: Exploratory changes in outcome measures

Across the six outcomes, estimated changes generally favoured KOKU-LITE; however, effect estimates were small and confidence intervals crossed zero, indicating considerable uncertainty. Positive standardised mean differences indicated change in favour of KOKU-LITE, whereas negative values indicated change in favour of control (Table 5).

**Table 5:**
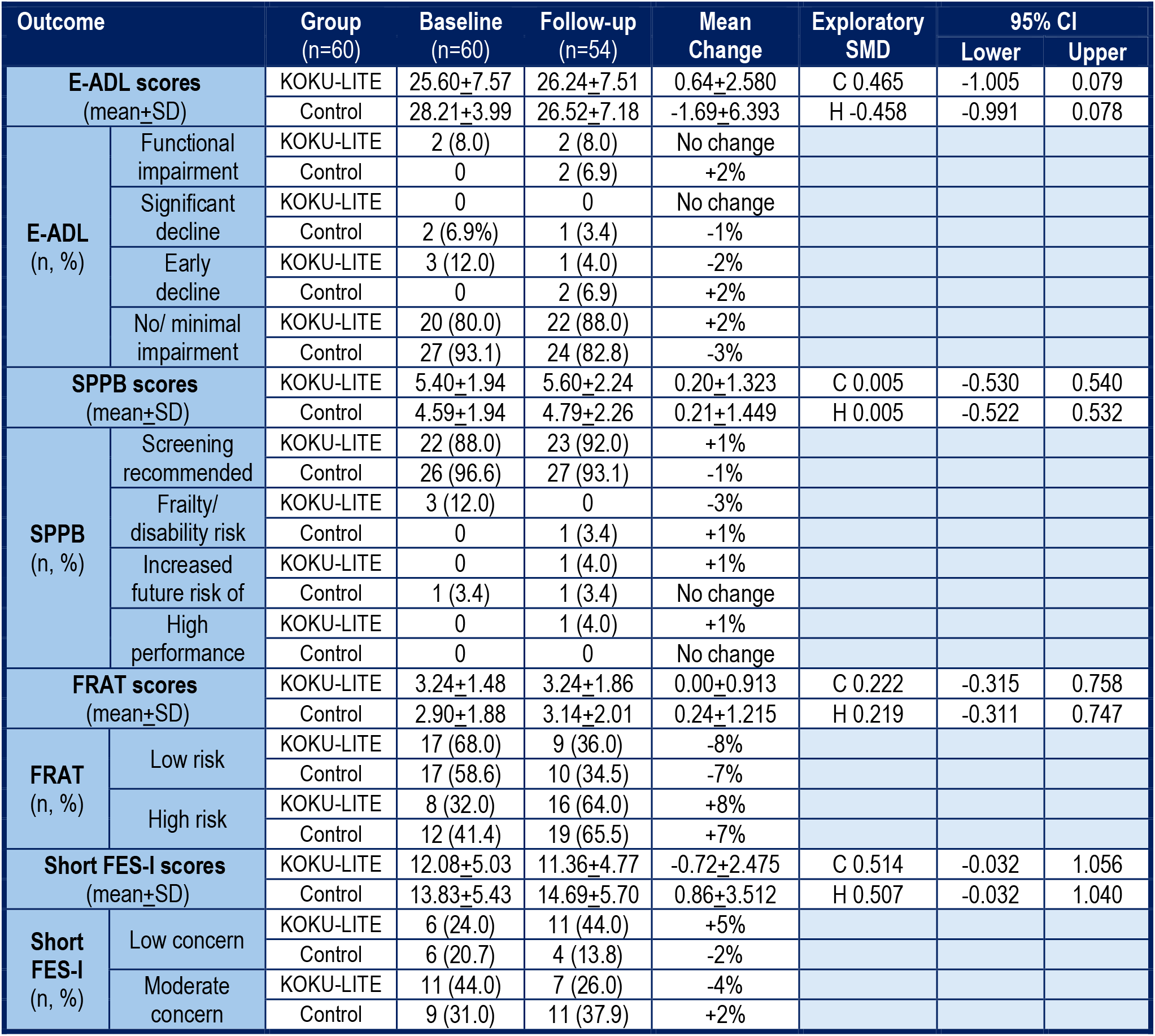
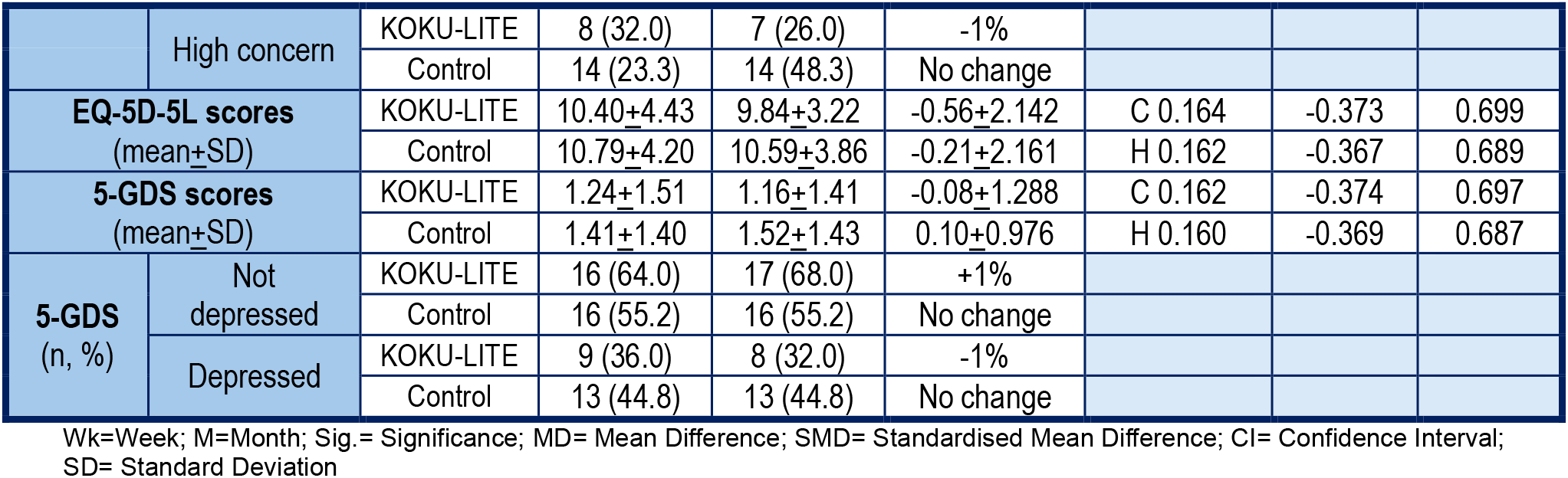
Pre-/post- test scores for the intervention and control groups.

The Short Falls Efficacy Scale - International (Short FES-I) scores [MD: −0.72; SMD: +0.514; CI: −0.032 to +1.056] demonstrated a moderate positive change favouring the intervention group, but as the CI crossed zero, this showed a trend in change, but not a statistically definitive change. The Erlangen Test for Activities of Daily Living (E-ADL) scores [MD: 0.64; SMD: +0.465; CI: −1.005 to +0.079] also showed a small to moderate, positive change towards the intervention group, but again due to the CI crossing zero, demonstrated no statistically significant difference and no clear evidence of real change in activities of daily living.

The Fall Risk Assessment Tool (FRAT) scores [MD: 0.00; SMD: +0.222; CI: −0.315 to +0.758] showed minimal change, with a very small trend slightly favouring the intervention with regards to falls risk, but this was statistically inconclusive. The 5-level European Quality of Life Healthy Questionnaire (EQ-5D-5L) scores [MD: −0.56; SMD: +0.164; CI: −0.373 to +0.699] showed a very small change in quality of life again slightly in favour of the intervention, but the change was small with no clear statistical difference. The 5-item Geriatric Depression Scale (5-GDS) scores [MD: −0.08; SMD: +0.162; CI: −0.374 to +0.697] showed a slight trend towards a reduction in depressive symptoms slightly in favour of the intervention group, but again the change was small and statistically inconclusive. Lastly the Short Physical Performance Battery (SPPB) scores showed a negligible difference in mean changes between groups [MD: 0.20; SMD: +0.005; CI: −0.53 to +0.54] indicating no meaningful between group difference in physical performance.

## 4. DISCUSSION

This feasibility randomised controlled trial investigated the acceptability, usability and feasibility of delivering KOKU-LITE, an evidence-based, dementia-friendly digital falls-prevention and wellbeing intervention, to PLwD in a home-based community setting. The study further examined the feasibility of recruitment and retention procedures, the appropriateness of candidate outcome measures, and the training and support requirements associated with future RCT implementation. Overall, the findings suggest that KOKU-LITE was acceptable to participants and feasible to deliver, and that the trial procedures were sufficiently robust to support progression to a larger, adequately powered definitive trial. Consistent with the objectives of a feasibility study, the trial was not designed to determine clinical effectiveness; therefore, the absence of statistically significant between group differences should not be interpreted as evidence of no intervention effect. Exploratory outcome trends were generally favourable to the intervention and, among the measures assessed, the Short FES-I and the E-ADL appeared most informative for a future trial, although the wide confidence intervals observed for functional outcomes indicate substantial uncertainty. FRAT, EQ-5D-5L, GDS-5 and SPPB demonstrated limited change over the six-week follow-up period and may be more appropriately retained as secondary or exploratory outcomes, or reconsidered in relation to follow-up duration, intervention dose and participant burden. The short follow-up period was pragmatic to inform the future RCT. Collectively, these findings support the feasibility of KOKU-LITE and provide evidence to inform refinement of the trial design, outcome selection and implementation strategy for a future definitive RCT.

These findings are consistent with an emerging body of evidence suggesting that digital, home-based exercise interventions may be acceptable to PLwD when intervention content is appropriately tailored, the technology is usable, and adequate technical support is available. Taylor et al.^(16)^ examined the feasibility of StandingTall, a tablet-based, individually tailored exercise programme delivered with caregiver assistance for community-dwelling PLwD. The study reported high levels of usability, enjoyment and adherence over 12 weeks. KOKU-LITE shares several key design and delivery features with StandingTall, including tablet-based delivery, home-based use and an emphasis on usability and acceptability. However, important differences should also be noted. StandingTall involved an older and smaller feasibility sample and required additional exercise equipment, whereas KOKU-LITE was evaluated in a larger feasibility sample and was designed as a simplified, dementia-adapted programme requiring minimal equipment. The Mais Equilibro (More Balance) study by Leite and colleagues^(32)^ provides supportive evidence for the usability and acceptability of app-based home exercises for falls prevention among older adults. However, because it was not developed or evaluated specifically for PLwD, its findings are only indirectly transferable to this population. KOKU-LITE therefore extends this evidence by providing dementia specific feasibility data, particularly in relation to usability, acceptability, support needs and implementation within a home-based community setting.

Evidence from non-digital and therapist supported exercise interventions further illustrates both the potential and the implementation challenges associated with falls prevention programmes for PLwD. The CogEx programme^(33)^ combined cognitive stimulation therapy with physical exercise in facilitated group sessions within aged care facilities. Although feasibility was demonstrated among people with mild-to-moderate cognitive impairment, challenges were identified in relation to recruitment, intervention fidelity and integration of cognitive and physical components. KOKU-LITE differs from CogEx in that it was designed for individual home-based use, which may reduce reliance on group-based delivery but increases the need for clear onboarding, accessible user support and remote monitoring. Suttanon et.al,^(34)^ reported improvements in balance and falls-risk outcomes following a structured home-based exercise programme for people with mild-to-moderate Alzheimer’s disease; however, the study was limited by its small sample size and reliance on supervised therapist visits. While such support may optimise safety and adherence, it also has implications for resource use and scalability. Whitney et al.^(35)^ evaluated ProF-Cog, a multifactorial intervention delivered in residential care with occupational therapist supervision, but reported limited clinical benefit and poor adherence, raising questions about feasibility in this setting. In contrast, Wesson et al.^(36)^ reported good feasibility, acceptability and adherence for a home hazard reduction and strength and balance intervention involving PLwD and carers, although no significant differences were observed in physiological outcomes. This pattern is broadly consistent with the present study, in which feasibility and acceptability are apparent, while evidence of clinical effectiveness remains to be established in a larger and longer definitive trial. The DIFRID mixed-methods study ^(37)^ further emphasises that home-based interventions for PLwD require careful adaptation, support and resource planning to enable successful delivery.

Collectively, the wider literature indicates that exercise-based interventions reduce falls or falls risk among older adults^(38)^ and may do so amongst PLwD^(39–41)^, although direct comparisons across studies remain limited by substantial methodological and clinical heterogeneity. Studies differ in participant characteristics, dementia subtype and severity, intervention content, mode of delivery, exercise dose and intensity, definitions of adherence, outcome measures and follow-up duration. These sources of heterogeneity are particularly important in dementia research, where cognitive symptoms, carer involvement, digital confidence, fatigue, comorbidity and safety considerations may influence both intervention engagement and outcome measurement. KOKU-LITE contributes to this evidence base by evaluating a dementia friendly gamified digital intervention in a community setting and by identifying feasible trial procedures and candidate outcomes for future evaluation. A definitive RCT should incorporate longer follow-up, more detailed monitoring of intervention exposure and engagement, clearer reporting of dementia related characteristics, and recruitment approaches that support the inclusion of underserved groups. These refinements are required before robust conclusions can be drawn regarding effectiveness, cost-effectiveness and implementation at scale.

This study has several strengths. It evaluated KOKU-LITE with PLwD in community-based home settings, providing directly relevant feasibility evidence for a population often underrepresented in digital falls prevention research. The sample was relatively large for a feasibility study in this field and included participants with different dementia subtypes and severity levels, supporting the planning and progression of a future definitive trial.

A further strength was the use of structured feasibility procedures and validated outcome measures. High retention and good tolerability of the assessments indicate that trial procedures were acceptable and feasible for this population, despite the recognised challenges of conducting exercise and digital health research with PLwD.

The findings should be interpreted in light of several limitations. The short intervention and follow-up period prevented assessment of sustained engagement or longer-term effects; however the study was designed as a feasibility study. Intervention exposure was not systematically monitored, limiting interpretation of dose, adherence and mechanisms of change. In addition, the sample was predominantly white British or Irish, which may limit transferability to more ethnically and culturally diverse populations. Future trials should therefore incorporate longer follow-up, robust engagement monitoring and recruitment strategies designed to improve inclusion of underserved groups using PROGRESS-Plus as an equity lens.

This study was not designed nor powered to determine clinical effectiveness, nonetheless these findings support progression to a definitive trial, but indicate several priorities for refinement. Future studies should use broader recruitment pathways, including clinical, community and voluntary sector routes specific to underserved communities.. Longer follow-up and systematic monitoring of intervention use are also required to assess adherence, intervention exposure and the long-term impact.

A future RCT should prioritise outcome measures that are sensitive to change, feasible for PLwD, and acceptable in terms of participant burden. In this study, the Short FES-I and the E-ADL appeared to be the most informative candidate secondary outcome measures. In addition, falls outcomes should be considered as the primary outcome to determine the effectiveness of KOKU-LITE. Consistent reporting of dementia subtype and severity will also be important to support interpretation and comparison across studies.

For implementation, KOKU-LITE suggests that simplified, home-based digital exercise interventions may be feasible for PLwD, but delivery at scale would require appropriate onboarding, technical support, carer involvement and attention to digital inclusion. Before adoption in routine dementia care, rehabilitation or falls-prevention pathways, adequately powered evidence is needed on clinical effectiveness, cost-effectiveness and acceptability across diverse populations.

KOKU-LITE was feasible to deliver and acceptable to PLwD in a home-based community setting, with feasible recruitment and retention procedures, well-tolerated outcome assessments, and no intervention-related serious adverse events. Although the study was not powered to assess effectiveness, the findings support progression to a definitive trial, with refinement of outcome selection, intervention use monitoring and follow-up duration.

A larger, adequately powered trial is required to determine whether KOKU-LITE improves falls-related outcomes, functional ability, confidence, quality of life and wellbeing, and whether it can be implemented equitably and cost-effectively at scale.

## Data Availability

Additional de-identified individual participant data will be available to the researchers upon request from the corresponding and senior author following completion of a Data Use Agreement.

## Contributors

Conceptualisation: JG, ES, EV; Ethical approval: JG, ES, EV; Methodology: JG, ES, EV; Study recruitment: JG, SA, AP; Data collection: JG, SA, AP; Data entry and cleaning: AA, SDD, SG, SW, RC, BK; Data analysis: SA, CS, SD, JG, RC; Writing original draft: JG, SA, ES

Writing, reviewing and editing: JG, SA, AP, CS, SDD, CT, EV, ES

## Data sharing

Additional de-identified individual participant data will be available to the researchers upon request from the corresponding and senior author following completion of a Data Use Agreement. The published protocol for the study can be found here:

Gangannagaripalli J, Vardy E, Stanmore E. A digital programme to prevent falls and improve well-being in people living with dementia in the community: the KOKU-LITE feasibility randomised controlled trial protocol. BMJ Open 2025;15:e091222. doi: 10.1136/bmjopen-2024-091222

## Acknowledgments

Thank you to the NIHR ARC-GM research assistants; Ms Bibhusha Karki, and NIHR ARC-GM interns; Ms Sheneil Goodman and Ms Stephanie Wynne, and the department summer intern Ms Azizat Amodu for their assistance with data entry, coding and cleaning for this feasibility study. Many thanks also to the project advisory group for their input and to the voluntary and community organisations for their help with recruiting participants and raising awareness of the study.

