## Supplementary appendix 2 for "A digital programme to prevent falls and improve wellbeing in people living with dementia in the community: Quantitative results from the KOKU-LITE feasibility study"

**Appendix 2: TIDieR reporting guideline checklist**

TIDieR
Template for Intervention Description and Replication

### The TIDieR (Template for Intervention Description and Replication) Checklist*:

Information to include when describing an intervention and the location of the information

| Item number | Item | Where located ** Primary paper (page or appendix number) | Other † (details) |
|---|---|---|---|
| | **BRIEF NAME** | | |
| 1. | Provide the name or a phrase that describes the intervention. | ____1__ | KOKU-LITE: A digital programme to prevent falls and improve wellbeing in people living with dementia in the community: Quantitative results from the KOKU-LITE feasibility study. |
| | **WHY** | | |
| 2. | Describe any rationale, theory, or goal of the elements essential to the intervention. | ____8,10________ | KOKU is a gamified, digital health programme designed to maintain function and reduce falls through strength & balance exercises (FaME/OTAGO), and health literacy games including hydration, nutrition, home safety, brain and bone health. KOKU has been modified to suit the needs and preferences of people living with dementia known as KOKULITE. This feasibility study has explored whether KOKU-LITE can be used to support people living with dementia in the community and to test the trial processes and its implementation. |
| | **WHAT** | | |
| 3. | Materials: Describe any physical or informational materials used in the intervention, including those provided to participants or used in intervention delivery or in training of intervention providers. Provide information | ______5,7,8______ | KOKU-LITE can be accessed from the KOKU Health website: https://kokuhealth.com/.<br>Intervention: KOKU-LITE + dementia specific falls prevention leaflet<br>Control: dementia specific falls prevention leaflet |

| | | | |
|---|---|---|---|
| | on where the materials can be accessed (e.g. online appendix, URL). | | dementia falls prevention leaflet can be accessed from: https://www.dementiauk.org/wp-content/uploads/dementia-uk-falls.pdf- |
| **4.** | Procedures: Describe each of the procedures, activities, and/or processes used in the intervention, including any enabling or support activities. | ______ 8________ | Over a 6-week period, participants were randomly allocated to either intervention (KOKU-LITE+dementia specific falls prevention leaflet) or the control (dementia specific falls prevention leaflet). Participants were be advised to use the KOKU-LITE for 30 minutes per session, 3 times per week, for 6 weeks along with the dementia specific falls prevention leaflet (for 6 weeks) and the control group participants to use dementia falls prevention leaflet for 6 weeks. |
| **5.** | **WHO PROVIDED**<br>For each category of intervention provider (e.g. psychologist, nursing assistant), describe their expertise, background and any specific training given. | ______8,9_______ | All balance/mobility assessments were completed by the trained and experienced researchers in the research team. Up to 30 minutes KOKU-LITE training was provided to participants by the trained and experienced researchers before the data collection commenced. |
| **6.** | **HOW**<br>Describe the modes of delivery (e.g. face-to-face or by some other mechanism, such as internet or telephone) of the intervention and whether it was provided individually or in a group. | ________8______ | Participants in the KOKU-LITE group attended a KOKU-LITE training session provided by trained and experienced researchers in the team lasting for up to 30 minutes. An iPad (with KOKU-LITE programme) was provided to participants for 6-weeks for their use at home, and they were advised to use KOKU-LITE for 30 minutes a day, 3 times per week, for 6 weeks. Participants in the control group were advised to use dementia specific falls prevention leaflet for 6 weeks. |

| | | | |
|---|---|---|---|
| **7.** | **WHERE**<br>Describe the type(s) of location(s) where the intervention occurred, including any necessary infrastructure or relevant features. | _____8________ | Intervention was delivered in participants' homes across Greater Manchester, UK. Participants were provided necessary training on how to use the KOKU-LITE programme and are advised to use KOKU-LITE programme at home. |
| **8.** | **WHEN and HOW MUCH**<br>Describe the number of times the intervention was delivered and over what period of time including the number of sessions, their schedule, and their duration, intensity or dose. | _______8______ | Participants were advised to use the KOKU-LITE for 30 minutes per session, 3 times per week, for 6 weeks along with the Dementia specific falls prevention leaflet. During the intervention period, the researcher provided technical assistance when required. |
| **9.** | **TAILORING**<br>If the intervention was planned to be personalised, titrated or adapted, then describe what, why, when, and how. | ______________ | The number of home visits to each participant differed based on their needs and preferences. The exercise programme in KOKU-LITE was tailored to each participant based on their abilities. |
| **10.‡** | **MODIFICATIONS**<br>If the intervention was modified during the course of the study, describe the changes (what, why, when, and how). | ___N/A___ | ______________ |
| **11.** | **HOW WELL**<br>Planned: If intervention adherence or fidelity was assessed, describe how and by whom, and if any strategies were used to maintain or improve fidelity, describe them. | ______8_______ | During the intervention period, researchers contacted participants by telephone to support adherence and troubleshoot technical issues. Intervention participants were also encouraged to read and use the dementia specific falls prevention leaflet. |

| | | | |
|---|---|---|---|
| **12.‡** | Actual: If intervention adherence or fidelity was assessed, describe the extent to which the intervention was delivered as planned. | ______________ | ______________ |

** **Authors** - use N/A if an item is not applicable for the intervention being described. **Reviewers** – use '?' if information about the element is not reported/not sufficiently reported.

† If the information is not provided in the primary paper, give details of where this information is available. This may include locations such as a published protocol or other published papers (provide citation details) or a website (provide the URL).

‡ If completing the TIDieR checklist for a protocol, these items are not relevant to the protocol and cannot be described until the study is complete.

* We strongly recommend using this checklist in conjunction with the TIDieR guide (see *BMJ* 2014;348:g1687) which contains an explanation and elaboration for each item.

* The focus of TIDieR is on reporting details of the intervention elements (and where relevant, comparison elements) of a study. Other elements and methodological features of studies are covered by other reporting statements and checklists and have not been duplicated as part of the TIDieR checklist. When a **randomised trial** is being reported, the TIDieR checklist should be used in conjunction with the CONSORT statement (see www.consort-statement.org) as an extension of **Item 5 of the CONSORT 2010 Statement.** When a **clinical trial protocol** is being reported, the TIDieR checklist should be used in conjunction with the SPIRIT statement as an extension of **Item 11 of the SPIRIT 2013 Statement** (see www.spirit-statement.org). For alternate study designs, TIDieR can be used in conjunction with the appropriate checklist for that study design (see www.equator-network.org).
